# Disease Risk Prediction Using Structured EHR Data: Can Generalist Large Language Models Match Specialized Clinical Foundation Models? A Comparative Evaluation with Fine-Tuning

**DOI:** 10.64898/2026.04.24.26351503

**Authors:** Bingyu Mao, Made K. Prasadha, Ziqian Xie, Jianping He, Michael Ghebranious, Hua Xu, Degui Zhi, Laila Rasmy

## Abstract

**Background:** Electronic health records (EHRs) with clinical decision support tools are now ubiquitous in healthcare organizations. Clinical foundation models (CFMs) pretrained on large-scale, heterogeneous structured EHR data have emerged as a powerful approach to improve predictive performance and generalizability. Meanwhile, large language models (LLMs) pretrained on broad data sources are being applied to an expanding range of healthcare tasks. However, it remains unclear whether generalist LLMs can match specialized CFMs for disease risk prediction using structured clinical data.

**Methods:** We compared CFMs (Med-BERT, CLMBR) against fine-tuned generalist LLMs (Mistral, LLaMA-2/3/3.1), a clinical LLM (Me-LLaMA), and LLM-generated embeddings paired with simple classifiers (using DeepSeek, Qwen3, and GPT-OSS) on two disease risk prediction tasks: heart failure risk among diabetic patients (DHF) and pancreatic cancer diagnosis (PaCa). Evaluations spanned multi-site EHR data, claims data, and an open-source single-institution benchmark (EHRSHOT). Performance was assessed using the area under the receiver operating characteristic curve (AUROC) and the area under the precision-recall curve (AUPRC).

**Results:** On larger EHR and claims cohorts (>30,000 patients), fine-tuned CFMs outperformed fine-tuned LLMs by a small but statistically significant margin (<1% AUROC). The clinical LLM performed comparably to generalist LLMs despite being smaller. On the open-source PaCa cohort (3,810 patients, 199 cases), LLMs achieved slightly higher AUROCs that were not statistically significant (LLaMA-3.1-70B 86.1% vs. Med-BERT 85.3%, p=0.27), but CFMs achieved significantly higher AUPRC (Med-BERT 55.9% vs. LLaMA-3.1-70B 41.1%, p=0.001). Notably, LLM-generated trajectory embeddings paired with logistic regression or a simple MLP, without any LLM fine-tuning, achieved the best overall performance, with AUROC exceeding 90% (Qwen3) and AUPRC reaching 66% (GPT-OSS 20B).

**Conclusion:** LLM-generated embeddings with lightweight classifiers outperformed both fine-tuned CFMs and fine-tuned LLMs on AUROC and AUPRC. While these results demonstrate the potential of generalist models to match or surpass specialized CFMs, their substantially greater computational cost and variable AUPRC performance in the fine-tuning setting warrant caution. We provide a reproducible evaluation framework and codebase to support continued benchmarking.

## 1. Introduction

Clinical decision support tools based on predictive models using structured electronic health records (EHRs) are commonly in use in healthcare organizations as they support early detection^1^, personalized treatment^2^, and resource optimization^2^. Over the past decade, machine learning-based predictive models have become the standard approach in clinical decision support. Pretrained models designed specifically for structured EHR data, such as Med-BERT^3^, BEHRT^4^, CLMBR^5^, TransformEHR^6^, and EHRMamba^7^, were developed mainly to improve the performance and the generalizability of such predictive models even with commonly known limitations of structured EHR data such as missingness, noise, inaccuracy, and sparsity. These models address these challenges primarily by generating contextual embeddings that represent the patient trajectory based on the parameters learned from complex patterns present in high-dimensional and longitudinal EHR data for millions of patients during pretraining.

Foundation models are defined as large AI models pretrained on broad data at scale to enable adaptability across a wide range of downstream tasks^8^. Following this terminology, we refer to pretrained models specifically designed for structured EHR data as clinical foundation models (CFMs), a class of specialized foundation models that have shown strong predictive performance on EHR-based tasks □. Meanwhile, generalist foundation models, especially large language models (LLMs), have disrupted many fields, including healthcare^10–12^, due to their adaptability and ability to generalize across diverse tasks. As awareness of these models’ capabilities grows, decision makers are increasingly exploring the use of LLMs for diagnostic decision-making^10–12^. This raises an intriguing question: Can LLMs outperform CFMs, boosting the performance of clinical predictive models using structured EHR data?

Evidence directly comparing LLMs to specialized CFMs remains limited. CFMs are explicitly designed and optimized for structured EHR data and have shown over 20% improvements in prediction accuracy^3^ and require less task-specific training data for fine-tuning when needed. On the other hand, generalist LLMs like GPT^13,14^ and LLaMA^15,16^, are pre-trained on various datasets and handle free-text inputs. Clinical LLMs (CLLMs) like Me-LLaMA^17^ are specialized versions of generalist LLMs, which are fine-tuned on domain-specific clinical text data, mainly clinical notes, also available in EHRs.

In the context of clinical prediction tasks, recent studies explored the potential of LLMs and compared their performance against specialized machine learning (ML) models. Chen et al^10^ and Hu et al.^18^ found that LLMs cannot beat traditional ML models in clinical prediction. CFMs have been shown to improve the performance of traditional ML models like logistic regression and deep learning models in many instances^3,5,9,19,20^. Traditional ML models may still perform well, particularly when trained on small local datasets, although they may tend to overfit. Shoham et al.^11^ and Acharya et al.^12^ fine-tuned LLaMA models on structured EHR data, reporting modest improvements over their respective versions of Med-BERT. Notably, these versions were pre-trained on a single source, such as MIMIC-IV, using relatively small cohorts compared to the scale typically required to pre-train an optimal CFM.

Unlike previous works, in this study we focused on comparing the utility of CFMs pre-trained on millions of patients’ full trajectories and LLMs for disease risk prediction, which commonly needs modeling for complex, long-term interactions within patient histories^3,11,12^. We restricted our evaluation to open-weight LLMs for two reasons: first, open-weight models allow supervised fine-tuning and embedding extraction on local infrastructure, which is essential for reproducibility; second, they avoid transmitting protected health information to external APIs, an important consideration for HIPAA compliance and institutional data governance in clinical settings. We compared the prediction discriminative accuracy of classifiers built on top of finetuned CFMs, such as Med-BERT and CLMBR^5^, versus fine-tuned LLMs such as Mistral^21^ and LLaMA^15,16^ and a CLLM Me-LLaMA^17^, for predicting patient risk of getting diagnosed with pancreatic cancer (PaCa) and the diabetic patients risk of developing heart failure (DHF) which using the same evaluation cohorts used in the Med-BERT^3^ paper as well as the PaCa cohort available in the EHRSHOT^19^ dataset. In this study, our contributions are summarized as follows:

1. We present the first head-to-head comparison of open-weight LLMs against CFMs pretrained at scale—including an encoder-based CFM trained on over 50 million patients’ longitudinal EHR and claims data (Med-BERT v2) and a decoder-based CFM trained on 2.57 million patients’ EHRs (CLMBR)—for disease risk prediction.
2. We evaluate the impact of different data serialization strategies for converting structured EHR data into textual input for LLMs.
3. We compare two modes of leveraging open-weight LLMs—supervised fine-tuning versus embedding extraction with lightweight classifiers—and show that the latter achieves the strongest overall performance.

## 2. Methods

### 2.1 Models

We evaluated three model categories: CFM, generalist LLM, and CLLM. CFMs are a specialized type of foundation model (FM) developed for electronic medical record (EMR) data, designed to address healthcare-specific tasks^9^. We tested two representative CFMs: Med-BERT and CLMBR. Med-BERT is an early encoder-based CFM trained on longitudinal structured EHR data from large-scale clinical datasets, and has been shown to substantially improve prediction accuracy for disease prediction tasks^3^. CLMBR is a decoder-transformer-based, autoregressive foundation model (CLMBR-T-base), originally described by Steinberg et al. (2021)^5^ and later improved by Wornow et al. (2023)^9^. For LLMs, five open-source generalist LLMs were selected for fine-tuning: the Mistral-7B-Instruct-v0.2^21^ model and LLaMA^15,16^ series models, including LLaMA-2-13b-hf, LLaMA-3-8B-Instruct, LLaMA-3.1-8B-Instruct, and LLaMA-3.1-70B-Instruct. Mistral-7B incorporates grouped-query attention (GQA)^22^ for faster inference by clustering related queries and sliding-window attention (SWA)^23^ to efficiently handle long sequences. LLaMA-2^15^ is an auto-regressive language model that features a standard Transformer architecture with extensions, including root mean square layer normalization (RMSNorm)^24^ for pre-normalization and a swish-gated linear unit (SwiGLU)^25^ activation function. It was fine-tuned using reinforcement learning from human feedback (RLHF)^26^. Finally, we included Me-LLaMA^17^, a CLLM built on LLaMA-2, which is continually pre-trained on 129B tokens and instruction-tuned with 214K clinical samples. Full model details and comparison of the key characteristics are provided in Appendices A and B of the supplementary materials. Additionally, as the LLM domain is fast-moving, we evaluated a group of the latest LLMs to directly generate patient representations, without fine-tuning, and trained a classifier on top of those pre-generated patient embeddings. For patient representations, we tested specialized LLMs such as MedGAMMA^27^ and Phi4^28^, and generalized LLMs with reasoning capabilities such as Deepseek^29^, GPT-OSS^30^, Qwen3^31^ thinking, and LLM embedding models such as Qwen3 8B.

### 2.2 Datasets and prediction tasks

This study focuses on two binary classification disease prediction tasks: DHF and PaCa, mirroring those addressed by Med-BERT^3^. The objective of these tasks is to predict whether a patient will be diagnosed with DHF or PaCa based on their structured EHR history. The prediction relies on patient history, including past diagnoses, medications, procedures, and other EHR information. To ensure a fair comparison, we used the same datasets evaluated in the Med-BERT study. These include two datasets derived from a structured EHR database (DHF-EHR and PaCa-EHR), and a patient-level claims dataset, PaCa-Claims. We also included the publicly available EHRSHOT dataset for the PaCa prediction task to further expand the evaluation. To explore the impact of data richness, we processed patient histories in three levels of clinical events categories: (1) diagnosis-only (D) includes only diagnosis information, which is the same as Med-BERT; (2) the combination of diagnosis, medication, and procedure (DMP), a richer representation similar to the Med-BERT v2^32^ preprocessing; and (3) all-codes (ALL), which incorporates all available structured data, such as demographics, vitals, laboratory results and clinical observations. Appendix C in the supplementary materials includes a detailed description of datasets and prediction tasks.

### 2.3 Data preprocessing

Structured EHR data were preprocessed to align with each model’s requirements. For Med-BERT, we followed the original preprocessing pipeline^3^, organizing patient visits chronologically and standardizing input lengths. For CLMBR, we used EHRSHOT scripts with minor modifications to ensure one prediction per patient and consistent prediction timing. LLM inputs were generated by converting structured data into text, following formats from CPLLM^11^ and LLaMA2-EHR^12^. We tested two prompt styles, the LLaMA2-EHR^12^ format with diagnostic frequency, to assess the impact on performance. For LLM embedding generation, due to run-time constraints, we generate embeddings for the first 4096 tokens of a patient’s history, including all visits and codes associated with each visit, ordered from most recent visit to oldest. Full preprocessing details are in Appendix D.

### 2.4 Fine-tuning and evaluation

All models were fine-tuned using the HuggingFace Transformers library. Med-BERT and CLMBR were pretrained on GPU clusters, with fine-tuning following a similar setup but with task-specific adjustments. LLMs and the CLLM were fine-tuned using parameter-efficient techniques, including low-rank adaptation of large language models (LoRA)^33^ from the parameter-efficient fine-tuning (PEFT)^34^ techniques and model quantization to reduce memory usage and training time.

Following the Med-BERT paper and other previous works on clinical prediction tasks^11,12^, we used the area under the receiver operating characteristic curve (AUROC) as the main evaluation metric for all experiments. For the DHF-EHR, PaCa-EHR, and PaCa-Claims tasks, which are the same as those in Med-BERT, we used diagnosis information as input, evaluated three traditional machine learning models: logistic regression (LR), random forest (RF), and light gradient boosting machine (LGBM). All five generalist LLMs and the CLLM were fine-tuned and evaluated on these three tasks. For the PaCa task on EHRSHOT, we evaluated three combinations of clinical events categories and included CLMBR, Med-BERT v2, the CLLM, and the two best-performing LLMs from the DHF-EHR, PaCa-EHR, and PaCa-Claims experiments. Full fine-tuning and evaluation details are provided in Appendices E and F, respectively.

### 2.6 Framework overview

Figure 1 shows the overview of the framework for each disease prediction task, as well as a summary of the results from selected experiments. Note that the patient information shown in the figure is not real patient data but just for illustration purposes. The process begins with the extraction of structured EHR data, which comprises both numerical medical codes and their corresponding textual descriptions. CFMs reformat the numerical codes following the data preprocessing methods of Med-BERT or CLMBR, while LLMs process the textual descriptions using prompt designs similar to CPLLM or LLaMA2-EHR. After preprocessing, both CFMs and LLMs are fine-tuned to generate predicted risk scores, which are subsequently evaluated using AUROC and AUPRC. Based on our findings, we also directly generate patient-level embeddings using different LLM models and file formats.

**Figure 1:**
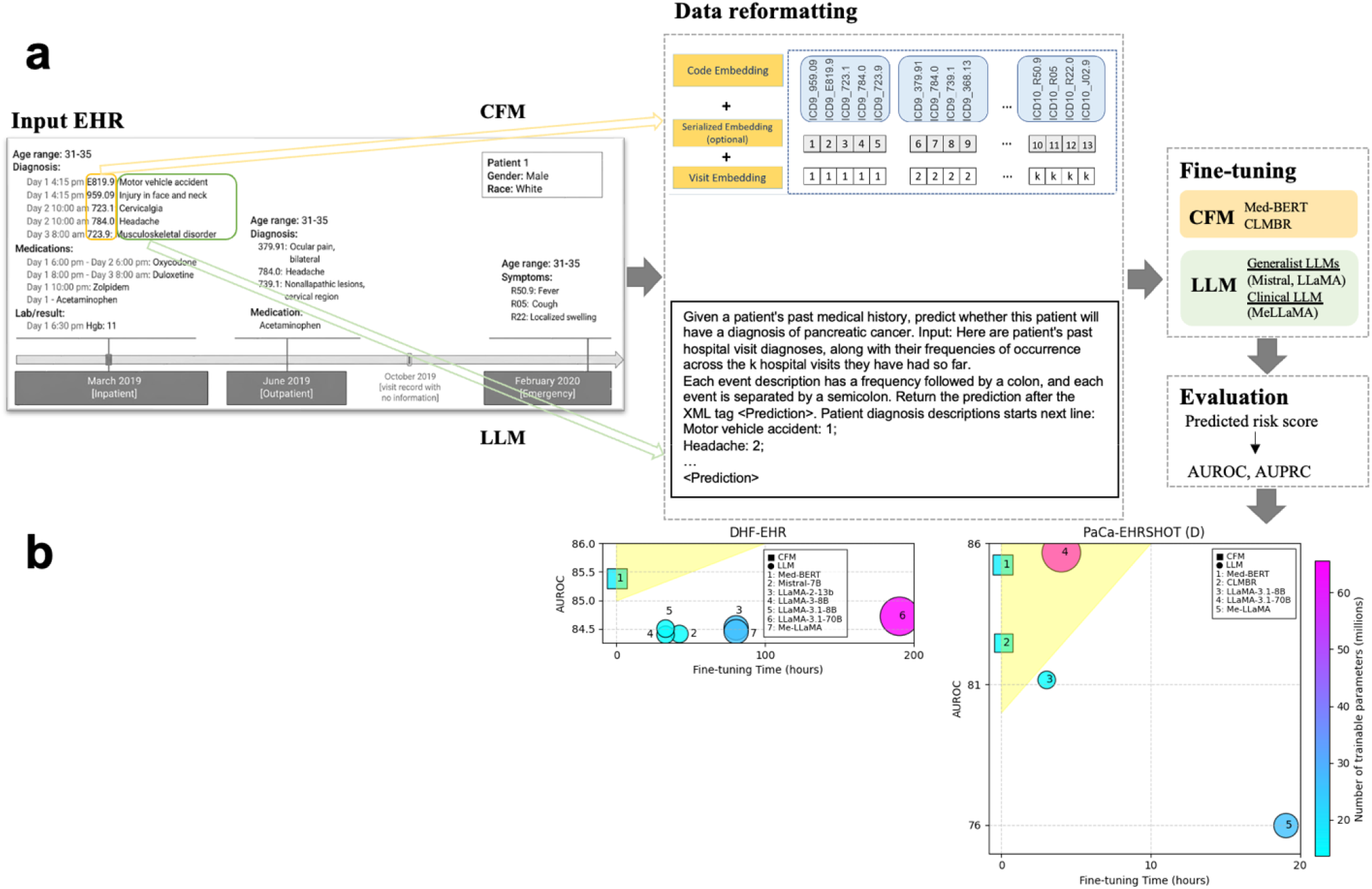
Project overview. (a) Methodology framework. (b) Main results. The patient information shown in this figure is not real patient data but just for illustration purposes.

A detailed code repository to reproduce our experiments can be found at: https://github.com/ClinicalFM/LLM2Predict.

## 3. Results

### 3.1 Descriptive analysis

Table 1 summarizes the descriptive analysis of the datasets and tasks included in this study, highlighting differences in patient sample size, visit frequency, and the scope of information used for modeling. Detailed descriptive analysis can be found in Appendix G of the supplementary materials.

**Table 1.**
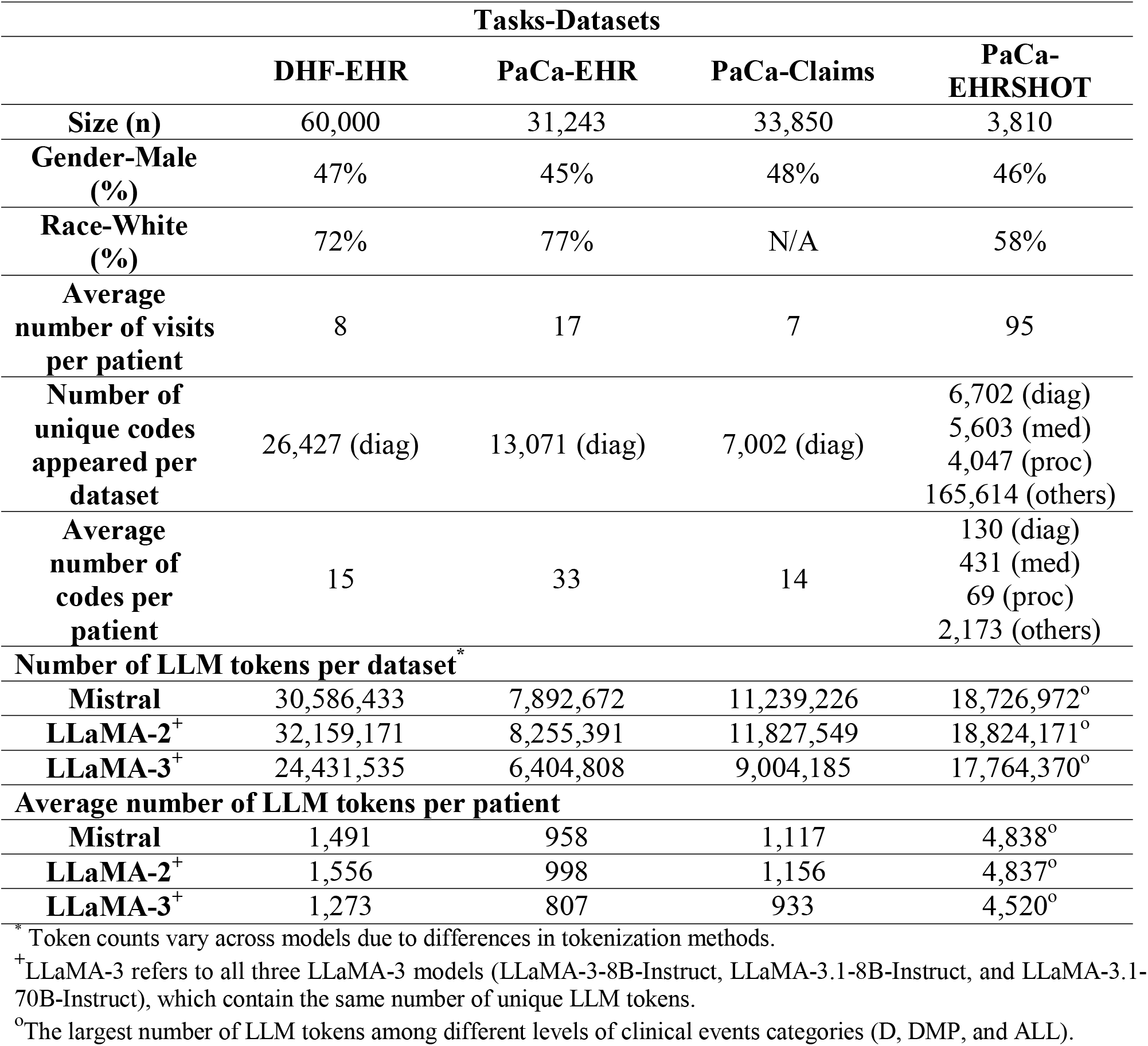
Descriptive analysis of the study cohort.

|  | Tasks-Datasets |  |  |  |
| --- | --- | --- | --- | --- |
|  | DHF-EHR | PaCa-EHR | PaCa-Claims | PaCa-EHRSHOT |
| **Size (n)** | 60,000 | 31,243 | 33,850 | 3,810 |
| **Gender-Male (%)** | 47% | 45% | 48% | 46% |
| **Race-White (%)** | 72% | 77% | N/A | 58% |
| **Average number of visits per patient** | 8 | 17 | 7 | 95 |
| **Number of unique codes appeared per dataset** | 26,427 (diag) | 13,071 (diag) | 7,002 (diag) | 6,702 (diag)<br>5,603 (med)<br>4,047 (proc)<br>165,614 (others) |
| **Average number of codes per patient** | 15 | 33 | 14 | 130 (diag)<br>431 (med)<br>69 (proc)<br>2,173 (others) |
| **Number of LLM tokens per dataset\*** |  |  |  |  |
| **Mistral** | 30,586,433 | 7,892,672 | 11,239,226 | 18,726,972<sup>o</sup> |
| **LLaMA-2<sup>+</sup>** | 32,159,171 | 8,255,391 | 11,827,549 | 18,824,171<sup>o</sup> |
| **LLaMA-3<sup>+</sup>** | 24,431,535 | 6,404,808 | 9,004,185 | 17,764,370<sup>o</sup> |
| **Average number of LLM tokens per patient** |  |  |  |  |
| **Mistral** | 1,491 | 958 | 1,117 | 4,838<sup>o</sup> |
| **LLaMA-2<sup>+</sup>** | 1,556 | 998 | 1,156 | 4,837<sup>o</sup> |
| **LLaMA-3<sup>+</sup>** | 1,273 | 807 | 933 | 4,520<sup>o</sup> |

\* Token counts vary across models due to differences in tokenization methods.
<sup>+</sup> LLaMA-3 refers to all three LLaMA-3 models (LLaMA-3-8B-Instruct, LLaMA-3.1-8B-Instruct, and LLaMA-3.1-70B-Instruct), which contain the same number of unique LLM tokens.
<sup>o</sup> The largest number of LLM tokens among different levels of clinical events categories (D, DMP, and ALL).

### 3.2 Fine-tuning results

Table 2 summarizes the average AUROC and standard deviations for different models across the DHF-EHR, PaCa-EHR, and PaCa-Claims tasks using the LLaMA2-EHR prompt format, offering important insights into model performance. Table S3 in Appendix H of the supplementary materials shows the model performance of tasks using the CPLLM prompt format, which consistently provides lower AUROCs than the LLaMA2-EHR format. The ML and CFM results in Table 2 are adapted from the results table of the Med-BERT paper^3^. Med-BERT + Bi-GRU achieved the highest AUROC (85.39) on DHF-EHR, while LLaMA-3.1-70B-Instruct and Me-LLaMA performed comparably (84.73 and 84.46). For PaCa-EHR, LLaMA-3.1-70B-Instruct and Me-LLaMA led with AUROCs of 82.96, slightly outperforming Med-BERT. Med-BERT remained best (80.57) in the PaCa-Claims task. P-values confirmed CFMs matched or outperformed LLMs significantly.

**Table 2.** Results table for tasks using the EHR and claims datasets.

|  |  | Average AUROC (STD) |  |  |
| --- | --- | --- | --- | --- |
| Models |  | DHF-EHR | PaCa-EHR | PaCa-Claims |
| ML | LR | 81.01 (0.00) | 79.94 (0.00) | 77.28 (0.00) |
|  | RF | 81.88 (0.08) | 79.48 (0.31) | 77.00 (0.12) |
|  | LGBM | 83.29 (0.00) | 81.78 (0.00) | 79.13 (0.00) |
|  | Bi-GRU | 82.82 (0.17) | 76.09 (0.61) | 76.79 (0.29) |
| CFM | Med-BERT | 85.18 (0.12) | 81.67 (0.31) | 79.98 (0.26) |
|  | Med-BERT + Bi-GRU | **85.39 (0.05)** | 82.23 (0.29) | **80.57 (0.21)** |
| LLM | Mistral-7B-Instruct-v0.2 | 84.42 (0.13) | 82.54 (0.15) | 79.39 (0.25) |
|  | LLaMA-2-13b-hf | 84.52 (0.08) | <u>82.70 (0.12)</u> | 79.68 (0.11) |
|  | LLaMA-3-8B-Instruct | 84.40 (0.15) | 82.32 (0.18) | 79.37 (0.17) |
|  | LLaMA-3.1-8B-Instruct | 84.51 (0.05) | 82.28 (0.08) | 79.71 (0.21) |
|  | LLaMA-3.1-70B-Instruct | <u>84.73 (0.20)</u> | **82.96 (0.16)** | 79.57 (0.54) |
| CLLM | Me-LLaMA | 84.46 (0.06) | **82.96 (0.30)** | <u>79.87 (0.15)</u> |
| P-value (the best CFM and the best LLM) |  | 0.0013 | 0.0024 | 0.00045 |

AUROC: the area under the receiver operating characteristics curve; STD: standard deviation for 5 repeats. Bold numbers are the best average AUROC (STD), and underlined numbers are the second-best ones.

Table 3 explores model performance for the PaCa-EHRSHOT task under three levels of clinical event categories: D (only include the diagnosis information), DMP (include diagnosis, medication, and procedure information), and ALL (include all available information). Since the original Med-BERT^3^ model was pre-trained only using the diagnosis information, we selected to test Med-BERT v2^32^, which was pre-trained on diagnosis, medication, and procedure information. Additionally, we only tested the better prompt format and the best generalist LLM based on the previous experiments, which are the LLaMA2-EHR prompt format and the LLaMA-3.1 series models. The results indicate that the LLaMA-3.1-70B-Instruct achieved the highest AUROCs in D (86.1) and DMP (86.65), while LGBM led in ALL (90.26). CFMs like Med-BERT v2 and CLMBR remained competitive to be the second best in D (85.25) and DMP (86.33). AUPRC results indicated that CFMs often outperformed LLMs, especially in D and DMP categories.

**Table 3.**
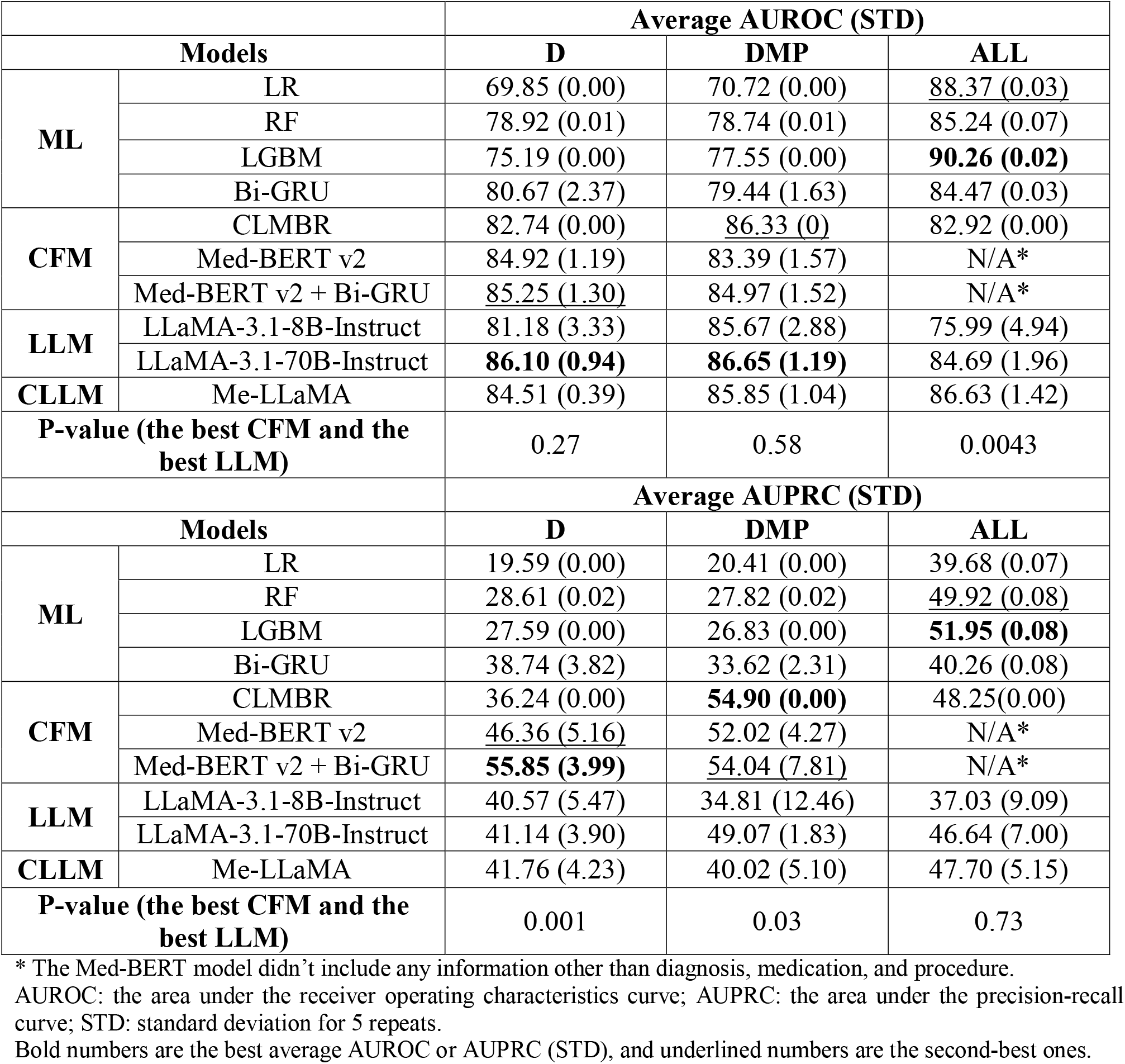
Results table for tasks using the EHRSHOT dataset.

|  |  | Average AUROC (STD) |  |  |
| --- | --- | --- | --- | --- |
| Models |  | D | DMP | ALL |
| ML | LR | 69.85 (0.00) | 70.72 (0.00) | <u>88.37 (0.03)</u> |
|  | RF | 78.92 (0.01) | 78.74 (0.01) | 85.24 (0.07) |
|  | LGBM | 75.19 (0.00) | 77.55 (0.00) | **90.26 (0.02)** |
|  | Bi-GRU | 80.67 (2.37) | 79.44 (1.63) | 84.47 (0.03) |
| CFM | CLMBR | 82.74 (0.00) | <u>86.33 (0)</u> | 82.92 (0.00) |
|  | Med-BERT v2 | 84.92 (1.19) | 83.39 (1.57) | N/A* |
|  | Med-BERT v2 + Bi-GRU | <u>85.25 (1.30)</u> | 84.97 (1.52) | N/A* |
| LLM | LLaMA-3.1-8B-Instruct | 81.18 (3.33) | 85.67 (2.88) | 75.99 (4.94) |
|  | LLaMA-3.1-70B-Instruct | **86.10 (0.94)** | **86.65 (1.19)** | 84.69 (1.96) |
| CLLM | Me-LLaMA | 84.51 (0.39) | 85.85 (1.04) | 86.63 (1.42) |
| P-value (the best CFM and the best LLM) |  | 0.27 | 0.58 | 0.0043 |
|  |  | Average AUPRC (STD) |  |  |
| Models |  | D | DMP | ALL |
| ML | LR | 19.59 (0.00) | 20.41 (0.00) | 39.68 (0.07) |
|  | RF | 28.61 (0.02) | 27.82 (0.02) | <u>49.92 (0.08)</u> |
|  | LGBM | 27.59 (0.00) | 26.83 (0.00) | **51.95 (0.08)** |
|  | Bi-GRU | 38.74 (3.82) | 33.62 (2.31) | 40.26 (0.08) |
| CFM | CLMBR | 36.24 (0.00) | **54.90 (0.00)** | 48.25(0.00) |
|  | Med-BERT v2 | <u>46.36 (5.16)</u> | 52.02 (4.27) | N/A* |
|  | Med-BERT v2 + Bi-GRU | **55.85 (3.99)** | <u>54.04 (7.81)</u> | N/A* |
| LLM | LLaMA-3.1-8B-Instruct | 40.57 (5.47) | 34.81 (12.46) | 37.03 (9.09) |
|  | LLaMA-3.1-70B-Instruct | 41.14 (3.90) | 49.07 (1.83) | 46.64 (7.00) |
| CLLM | Me-LLaMA | 41.76 (4.23) | 40.02 (5.10) | 47.70 (5.15) |
| P-value (the best CFM and the best LLM) |  | 0.001 | 0.03 | 0.73 |

\* The Med-BERT model didn't include any information other than diagnosis, medication, and procedure.
AUROC: the area under the receiver operating characteristics curve; AUPRC: the area under the precision-recall curve; STD: standard deviation for 5 repeats.
Bold numbers are the best average AUROC or AUPRC (STD), and underlined numbers are the second-best ones.

Moreover, Tables S4 and S5 in Appendix I of the supplementary materials compare the fine-tuning process speed by hours for all generalist and clinical LLMs. Mistral-7B was fastest among LLMs (42 hours), while LLaMA-3.1-70B required up to 190 hours. In contrast, CFMs completed fine-tuning in under 1 hour. The results summary in Figure 1 shows the results from DHF-EHR and PaCa-EHRSHOT (D), including the comparisons of AUROC, fine-tuning time, and number of trainable parameters for different models. The yellow area at the upper left corner of each subplot represents the models given the highest AUROC with the lowest fine-tuning time.

Table 4 shows that we achieve the best pancreatic cancer prediction performance using LLMs as encoders, without the need for fine-tuning. We tested various machine learning and basic MLP prediction heads and found that a simple logistic regression (AUROC 79.9% - 89.9%) or MLP (78.8% - 87.1%) yielded the best performance compared to tree-based (66.9% - 82.9%). We also found that adding instructions was commonly associated with better performance (67.7% - 89.9%) compared to without instructions (66.9% - 88.5%), and that using Markdown with a patient summary at the top was more effective (70.3% - 89.9%) compared to using the patient trajectory expanded in XML format (66.9% - 85.2%) (Supplementary Table 7). Additionally, we found that the latest embedding models, such as Qwen3, even with a lower number of parameters (8B) (85.1% - 90.8%), showed better performance than other instruction and thinking variant models with slightly larger parameters (20B) (82.8% - 88.4%). These results demonstrate that newer open-source models can efficiently generate helpful patient representations that, with a simple logistic regression model on top of it, get strong PaCa prediction results.

**Table 4.** Performance of LLMs as encoders for pancreatic cancer prediction in the EHRSHOT all codes cohort.

| Model | LR |  | MLP |  |
| --- | --- | --- | --- | --- |
|  | AUROC (STD) | AUPRC (STD) | AUROC (STD) | AUPRC (STD) |
| DeepSeek R1 70B + Markdown | <u>89.9</u> (00.0) | 59.9 (00.0) | 87.1 (0.500) | 53.9 (3.40) |
| GPT OSS 20B+ Markdown | 88.4 (00.0) | **66.1 (00.0)** | 86.6 (0.400) | 54 (3.5) |
| Llama 3.1 70B+ Markdown | 88.5 (00.0) | 59.9 (00.0) | 86.6 (0.700) | 51.7 (3.60) |
| MedGemma 27B+ Markdown | 86.9 (00.0) | 59.4 (00.0) | 87.2 (0.600) | 57.9 (1.80) |
| MedGemma 4B+ Markdown | 85.8 (00.0) | 54.9 (00.0) | 86 (0.200) | 48.9 (2.00) |
| Phi 4+ Markdown | 84.6 (00.0) | 46.3 (00.0) | 85.1 (0.500) | 46 (2.10) |
| Qwen2 7B + Markdown | 84.9 (00.0) | 53.4 (00.0) | 86 (1.30) | 53.6 (3.70) |
| Qwen3 8B Embedding + Text | 89.1 (00.0) | 56.4 (00.0) | **90.8 (0.436)** | <u>64.7 (1.98)</u> |
| Qwen3 8B Embedding + Markdown | 85.1 (00.0) | 55.8 (00.0) | 88.1 (0.200) | 52.2 (4.70) |
| Qwen3 30B Thinking + Text | 88.2 (00.0) | 50.2 (00.0) | 86.0 (1.00) | 34.4 (2.30) |
| Qwen3 30B Thinking + Markdown | 86.5 (00.0) | 53.8 (00.0) | 86.4 (0.500) | 46.6 (3.20) |
| Qwen3 30B Instruct + Text | 88.0 (00.0) | 47.9 (00.0) | 86.6 (1.20) | 38.1 (3.20) |
| Qwen3 30B Instruct + Markdown | 86.5 (00.0) | 54.0 (00.0) | 86.4 (0.600) | 47.6 (2.60) |

AUROC: the area under the receiver operating characteristics curve; AUPRC: the area under the precision-recall curve; STD: standard deviation for 5 repeats.
Bold numbers are the best average AUROC or AUPRC, and underlined numbers are the second-best ones.

## 4. Discussion

In this study, we presented a comparative evaluation of generalist LLMs and specialized CFMs for disease risk prediction using structured EHR data from different real-world sources. We focused on two disease risk prediction tasks, Heart Failure for diabetic patients and Pancreatic Cancer, as our use cases, across three datasets from different sources: (1) EHR data from multiple health systems across the US (DHF–EHR, PaCa-EHR), (2) claims data (PaCa-Claims), and (3) an open-source cohort (PaCa-EHRSHOT), to support reproducibility and future benchmarking. We also tested three different combinations of clinical event categories on EHRSHOT, i.e., diagnosis codes only (D), diagnosis, medications, and procedures combination (DMP), and all codes available in the EHRSHOT dataset (ALL), including demographics, vitals, laboratory results, and clinical observations in addition to the previously mentioned clinical event types. While LLMs show promising results, CFMs and, sometimes, even simple ML models achieve slightly better discriminative accuracy, especially when fine-tuned on larger cohorts. For example, Med-BERT reached an AUROC of 85.4 for DHF prediction, outperforming LLaMA-3.1-70B (AUROC=84.7). Additionally, CFMs achieved significantly higher AUPRCs than LLMs in all EHRSHOT experiments, even when LLMs showed slightly better AUROC. However, when utilizing LLMs to generate patient-level embeddings for a text summary of their full trajectory, they lead to the best performance (for example, PaCa-EHRSHOT-ALL: AUROC of 90.8 and AUPRC of 64.7 using a 3-layer MLP on top of Qwen3 embeddings). Compared to previous studies that mostly compare LLMs to ML models^10–12,18,35^, our work contributes to benchmarking against specialized CFMs trained on tens of millions of patients’ data, such as Med-BERT, for disease risk prediction, while also highlighting how data representation, patient information formats, and LLM utilization modes influence model performance.

From our experiments, we observed several key findings. First, converting patient trajectories into text using the LLaMA2-EHR^12^ format led to approximately 1% higher discriminative accuracy compared to the CPLLM^11^ format (Supplementary Appendix H) and accordingl,y we followed the LLaMA2-EHR ^12^ format for all experiments reported in Tables 2 and 3. This improvement may be partly due to the inclusion of additional information such as event counts. We also noted that differences in token granularity, e.g. SentencePiece^15^ used in LLaMA-2 versus Tiktoken^16^ in LLaMA-3, are among the factors impacting the variability in performance as well as computational resource needs. Second, we found that in the majority of our experiments except for PaCa-Claims, LLaMA-3.1-70B is achieving the best results among all other LLMs tested, given it was the largest model we fine-tuned in this study. However, the difference was just marginal (<0.3%). Third, we found that the CLLM (Me-LLaMA), which was further trained on clinical text, performed comparably to generalist LLMs. For example, Me-LLaMA is leading to around 83% AUROC for the PaCa-EHR experiment which is the same as the LLaMA-3.1-70B model performance, even though it is a smaller model (13B). Med-BERT achieved the highest AUPRC (55.9%) on the PaCa-EHRSHOT (D) task, outperforming CLMBR (DMP) (54.9%) and LGBM (ALL) (52.0%). In contrast, the best-performing LLM (LLaMA3.1-70B on DMP) reached only 49.1%. This may be partly attributed to the quality of terminology mapping: medication codes mapping from RxNorm to Multum identifiers is nearly complete (90%), followed by mapping the diagnosis codes between standard OMOP SNOMED codes to ICD codes (82%), but the mapping of the procedure codes was just at 10%. Notably, Med-BERT v2 was trained only on diagnosis, medication, and procedure codes, and thus was excluded from the all-codes experiments (Table 3). Similarly, as we could not map a high percentage (>60%) of our multi-site EHR or claims data to the CLMBR vocabulary, we decided not to report the CLMBR model performance in Table 2. Lastly, while we only used LORA adaptors with the default configuration during the LLMs’ supervised fine-tuning (SFT) in this study’s experiments for computational efficiency, the SFT of the LLM models was at least 40x longer than the SFT of CFM and it required at least double the computational resources. For example, for PaCa-EHRSHOT which is our smallest cohort, the fine-tuning of Med-BERT and CLMBR took less than 5 minutes using a single GPU, while LLaMA2-3.1-8B (13M tunable parameters) took around 4 hours using 2 H100 80GB GPUs on the DMP experiment, Me-LLaMA (13B with 26M tunable parameters) took around 7 hours, and LLaMA2-3.1-70B (65M) took 18 hours. Nonetheless, the LLaMA2-3.1-70B (ALL) took 110 hours using 4 GPUs (Supplementary Appendix I).

While CFM is currently performing better on disease risk prediction tasks using longitudinal structured EHR data given the data sources, modalities, and pretraining tasks used to train such models, generalist LLMs have proven ability to outperform on many clinical tasks, such as question answering, clinical note summarization, automatic medical coding, and clinical trial matching^36^. Recent advances further demonstrate their potential: Health-LLM^37^ achieved strong multimodal health prediction by combining contextual and physiological data, reaching performance comparable to larger models like GPT-3.5 and GPT-4; prompt engineering and retrieval-augmented generation (RAG) can further enhance their performance on medical tasks^38–40^. Additionally, studies such as Acharya et al.^12^ and Beaulieu-Jones et al.^35^, showed that LLM models fine-tuned on clinical data can outperform traditional ML models for clinical prediction tasks. However, studies such as Chen et al. (2024)^10^ and Brown et al. (2025)^41^ have demonstrated the opposite. We are one of the early studies comparing LLMs to CFMs trained on millions of patients records including Med-BERT v2 trained on more than 50 million patients data, and our findings echo Hegselmann et al. (2025)^42^, showing that LLMs can achieve comparative performance to specialized models. They compared LLM-embedding models (GTE-Qwen2-7B-Instruct and LLM2Vec-Llama3.1-8B-Instruct) versus CLMBR on the 15 clinical prediction tasks from the EHRSHOT benchmark and UK Biobank, and found that LLM encoders can outperform CLMBR in disease onset prediction, although they did not specify the performance on PaCa cohort. Our findings demonstrate the promise of the use of LLMs for disease prediction. However, instead of completely replacing CFMs, future work should consider integrating those models to get the advantage from both. For example, LLM-based embeddings can be utilized while training CFMs’ to further enhance their performance and generalizability as proposed by Su et al. (2025)^43^.

However, this study is not without limitations. First, our evaluation is limited to binary classification-based disease prediction tasks, and it remains unclear whether the observed trends extend to multi-class or regression-based clinical predictions. Second, while we used multiple datasets, the scope of patient information was limited, and including a wider range of clinical features could potentially improve model performance. Third, the models evaluated in this study are based on structured EHR data, and their performance on unstructured data, such as clinical notes, remains unexplored. Future work could expand on this by evaluating the impact of context length on model performance, as explored by Wornow et al. (2025)^44^, or by investigating hybrid approaches that combine CFMs and LLMs, as proposed by Hegselmann et al. (2025)^42^. In conclusion, our study has shown that generalist LLMs have the promise to improve the performance of disease prediction tasks. However, there are a lot of aspects that need to be considered before fully relying on them, that includes how the data will be presented to the LLM, key parameters considered during the LLM fine-tuning, computational resources available, data privacy and HIPAA compliance especially for open-source models, and the main metrics crucial to evaluate the clinical utility of the fine-tuned model. While that highlights the idea that LLMs are an exciting new direction in clinical prediction, CFMs continue to play an important role in healthcare and should not be overlooked. Future research should continue to refine the application of LLMs in clinical settings and explore ways to integrate the strengths of both LLMs and CFMs for improved disease prediction.

## Data Availability

All data produced in the present study are available upon reasonable request to the authors.

https://som-shahlab.github.io/ehrshot-website/

## Institutional Review Board Statement

All data used in this study were subject to oversight and approval by the Committee for the Protection of Human Subjects (UTHSC-H IRB) under protocol HSC-SBMI-13-0549.

## Data and Code Availability

For reproducibility, our codebase and the trained models will be available at https://github.com/ClinicalFM/LLM2Predict; however, for patient data privacy, confidentiality, and ethical reasons, the model training data cannot be shared. However, to reproduce results on the EHRshot cohort, access can be requested through https://som-shahlab.github.io/ehrshot-website/.

## Funding Acknowledgments

This study received funding in part from the National Institutes of Health and the National Library of Medicine grant R01LM014249.

## Authors contribution

BM led the study, contributed to study design and LLM fine-tuning, and drafted the manuscript. MKP led training of classifiers on LLM embeddings and contributed to manuscript drafting. ZX contributed to study design and analysis. MG contributed to data acquisition and preparation and provided CLMBR and MedBERT v2 results for the EHRshot cohort. JH fine-tuned the MeLLAMA model and provided evaluation results. HX contributed to study design, interpretation, and critical revision of the manuscript. DZ co-supervised the project with LR and contributed to study conception, analysis, interpretation, drafting, and critical revision of the manuscript. LR supervised the overall project and contributed to study conception and design, data acquisition, analysis, interpretation, drafting, and critical revision of the manuscript. All authors critically reviewed the manuscript, approved the final version, and agreed to be accountable for all aspects of the work.

## References

1. Topol EJ. Medical forecasting. Science 2024;384(6698):eadp7977. DOI: 10.1126/science.adp7977.

2. Topol EJ. High-performance medicine: the convergence of human and artificial intelligence. Nat Med 2019;25(1):44–56. DOI: 10.1038/s41591-018-0300-7.

3. Rasmy L, Xiang Y, Xie Z, Tao C, Zhi D. Med-BERT: pretrained contextualized embeddings on large-scale structured electronic health records for disease prediction. Npj Digit Med 2021;4(1):1–13. DOI: 10.1038/s41746-021-00455-y.

4. Li Y, Rao S, Solares JRA, et al. BEHRT: Transformer for Electronic Health Records. Sci Rep 2020;10(1):7155. DOI: 10.1038/s41598-020-62922-y.

5. Steinberg E, Jung K, Fries JA, Corbin CK, Pfohl SR, Shah NH. Language models are an effective representation learning technique for electronic health record data. J Biomed Inform 2021;113:103637. DOI: 10.1016/j.jbi.2020.103637.

6. Yang Z, Mitra A, Liu W, Berlowitz D, Yu H. TransformEHR: transformer-based encoder-decoder generative model to enhance prediction of disease outcomes using electronic health records. Nat Commun 2023;14(1):7857. DOI: 10.1038/s41467-023-43715-z.

7. Fallahpour A, Alinoori M, Ye W, Cao X, Afkanpour A, Krishnan A. EHRMamba: Towards Generalizable and Scalable Foundation Models for Electronic Health Records. In: Proceedings of the 4th Machine Learning for Health Symposium. Vancouver, Canada: PMLR, 2025: 291–307. (https://proceedings.mlr.press/v259/fallahpour25a.html)

8. Bommasani R, Hudson DA, Adeli E, et al. On the Opportunities and Risks of Foundation Models. 2022. (http://arxiv.org/abs/2108.07258)

9. Wornow M, Xu Y, Thapa R, et al. The shaky foundations of large language models and foundation models for electronic health records. Npj Digit Med 2023;6(1):1–10. DOI: 10.1038/s41746-023-00879-8.

10. Chen C, Yu J, Chen S, et al. ClinicalBench: Can LLMs Beat Traditional ML Models in Clinical Prediction? 2024. (http://arxiv.org/abs/2411.06469)

11. Shoham OB, Rappoport N. CPLLM: Clinical prediction with large language models. PLOS Digit Health 2024;3(12):e0000680. DOI: 10.1371/journal.pdig.0000680.

12. Acharya A, Shrestha S, Chen A, et al. Clinical risk prediction using language models: benefits and considerations. J Am Med Inform Assoc 2024;ocae030. DOI: 10.1093/jamia/ocae030.

13. Radford A, Narasimhan K, Salimans T, Sutskever I. Improving Language Understanding by Generative Pre-Training. 2018. (https://openai.com/research/language-unsupervised)

14. Brown T, Mann B, Ryder N, et al. Language Models are Few-Shot Learners. In: Proceedings of the Neural Information Processing Systems Conference. NeurIPS, 2020: 1877–1901. (https://papers.nips.cc/paper/2020/hash/1457c0d6bfcb4967418bfb8ac142f64a-Abstract.html)

15. Touvron H, Martin L, Stone K, et al. Llama 2: Open Foundation and Fine-Tuned Chat Models. 2023. (https://arxiv.org/abs/2307.09288v2)

16. Dubey A, Jauhri A, Pandey A, et al. The Llama 3 Herd of Models. 2024. (https://arxiv.org/abs/2407.21783v2)

17. Xie Q, Chen Q, Chen A, et al. Medical foundation large language models for comprehensive text analysis and beyond. Npj Digit Med 2025;8(1):141. DOI: 10.1038/s41746-025-01533-1.

18. Hu Y, Zuo X, Zhou Y, et al. Information extraction from clinical notes: are we ready to switch to large language models? J Am Med Inform Assoc 2026;33(3):553–562. DOI: 10.1093/jamia/ocaf213.

19. Wornow M, Thapa R, Steinberg E, Fries JA, Shah N. EHRSHOT: An EHR Benchmark for Few-Shot Evaluation of Foundation Models. Poster presented at Neural Information Processing Systems, New Orleans, LA, Dec 10–16, 2023. (https://bytez.com/docs/neurips/73656/paper)

20. Steinberg E, Fries JA, Xu Y, Shah N. MOTOR: A Time-to-Event Foundation Model For Structured Medical Records. In: Proceedings of the Twelfth International Conference on Learning Representations. Vienna, Austria: ICLR, 2024. (https://openreview.net/forum?id=NialiwI2V6)

21. Jiang AQ, Sablayrolles A, Mensch A, et al. Mistral 7B. 2023. (https://arxiv.org/abs/2310.06825v1)

22. Ainslie J, Lee-Thorp J, Jong M de, Zemlyanskiy Y, Lebron F, Sanghai S. GQA: Training Generalized Multi-Query Transformer Models from Multi-Head Checkpoints. In: Proceedings of the 2023 Conference on Empirical Methods in Natural Language Processing. Singapore: EMNLP, 2023: 4895–4901. (https://openreview.net/forum?id=hmOwOZWzYE)

23. Beltagy I, Peters ME, Cohan A. Longformer: The Long-Document Transformer. 2020. (http://arxiv.org/abs/2004.05150)

24. Zhang B, Sennrich R. Root mean square layer normalization. In: Proceedings of the 33rd International Conference on Neural Information Processing Systems. Vancouver, Canada: NeurIPS, 2019: 12381–12392. (https://dl.acm.org/doi/10.5555/3454287.3455397)

25. Shazeer N. GLU Variants Improve Transformer. 2020. (http://arxiv.org/abs/2002.05202)

26. Kaufmann T, Weng P, Bengs V, Hüllermeier E. A Survey of Reinforcement Learning from Human Feedback. Trans Mach Learn Res 2025. (https://openreview.net/forum?id=f7OkIurx4b)

27. Sellergren A, Kazemzadeh S, Jaroensri T, et al. MedGemma Technical Report. 2025. (http://arxiv.org/abs/2507.05201)

28. Abdin M, Aneja J, Behl H, et al. Phi-4 Technical Report. 2024. (http://arxiv.org/abs/2412.08905)

29. Guo D, Yang D, Zhang H, et al. DeepSeek-R1 incentivizes reasoning in LLMs through reinforcement learning. Nature 2025;645(8081):633–638. DOI: 10.1038/s41586-025-09422-z.

30. OpenAI, Agarwal S, Ahmad L, et al. gpt-oss-120b & gpt-oss-20b Model Card. 2025. (http://arxiv.org/abs/2508.10925)

31. Yang A, Li A, Yang B, et al. Qwen3 Technical Report. 2025. (http://arxiv.org/abs/2505.09388)

32. Rasmy L, Chu Y, Mao B, et al. Med-BERT v2: clinical foundation model on standardized secondary clinical data. In: Proceedings of the Machine Learning For Healthcare Conference. Durham, NC: MLHC, 2022. (https://www.researchgate.net/publication/361569724_Med-ERT_v2_clinical_foundation_model_on_standardized_secondary_clinical_data)

33. Hu EJ, Shen Y, Wallis P, et al. LoRA: Low-Rank Adaptation of Large Language Models. In: Proceedings of the Tenth International Conference on Learning Representations. ICLR, 2022. (https://openreview.net/forum?id=nZeVKeeFYf9)

34. Xu L, Xie H, Qin SJ, Tao X, Wang FL. Parameter-Efficient Fine-Tuning Methods for Pretrained Language Models: A Critical Review and Assessment. IEEE Trans Pattern Anal Mach Intell 2026;(01):1–20. DOI: 10.1109/TPAMI.2026.3657354.

35. Beaulieu-Jones BK, Villamar MF, Scordis P, et al. Predicting seizure recurrence after an initial seizure-like episode from routine clinical notes using large language models: a retrospective cohort study. Lancet Digit Health 2023;5(12):e882–94. DOI: 10.1016/S2589-7500(23)00179-6.

36. Wang Z, Wang H, Danek B, et al. A perspective for adapting generalist AI to specialized medical AI applications and their challenges. Npj Digit Med 2025;8(1):429. DOI: 10.1038/s41746-025-01789-7.

37. Jin M, Yu Q, Zhang C, et al. Health-LLM: Personalized Retrieval-Augmented Disease Prediction Model. 2024. (http://arxiv.org/abs/2402.00746)

38. Wang L, Chen X, Deng X, et al. Prompt engineering in consistency and reliability with the evidence-based guideline for LLMs. NPJ Digit Med 2024;7:41. DOI: 10.1038/s41746-024-01029-4.

39. Nori H, Lee YT, Zhang S, et al. Can Generalist Foundation Models Outcompete Special-Purpose Tuning? Case Study in Medicine. 2023. (http://arxiv.org/abs/2311.16452)

40. Xiong G, Jin Q, Lu Z, Zhang A. Benchmarking Retrieval-Augmented Generation for Medicine. In: Proceedings of Findings of the Association for Computational Linguistics. Bangkok, Thailand: ACL, 2024: 6233–6351. (https://aclanthology.org/2024.findings-acl.372/)

41. Brown KE, Yan C, Li Z, et al. Large language models are less effective at clinical prediction tasks than locally trained machine learning models. J Am Med Inform Assoc 2025;32(5):811–22. DOI: 10.1093/jamia/ocaf038.

42. Hegselmann S, Arnim G von, Rheude T, et al. Large Language Models are Powerful EHR Encoders. 2025. (http://arxiv.org/abs/2502.17403)

43. Su X, Messica S, Huang Y, et al. Multimodal Medical Code Tokenizer. Poster presented at the Forty-Second International Conference on Machine Learning, Vancouver, Canada, Jul 13–19, 2025. (https://openreview.net/forum?id=UaTrcei5Ba)

44. Wornow M, Bedi S, Hernandez MAF, et al. Context Clues: Evaluating Long Context Models for Clinical Prediction Tasks on EHRs. 2025. (http://arxiv.org/abs/2412.16178)

