## Supplementary materials for "Disease Risk Prediction Using Structured EHR Data: Can Generalist Large Language Models Match Specialized Clinical Foundation Models? A Comparative Evaluation with Fine-Tuning"

### Appendix A Model description

#### A1 Clinical Foundation Models (CFM)

CFMs are a specialized type of foundation model (FM) developed for electronic medical record (EMR) data, designed to address healthcare-specific tasks^9^. Like general FMs, CFMs are pre-trained on large amounts of unlabeled data using self-supervised learning to identify patterns and representations applicable across a wide variety of downstream tasks. However, CFMs are designed for clinical contexts and can be broadly categorized into two subtypes, clinical language models such as Me-LLaMA^17^ trained mostly on clinical text and foundation models trained on structured real-world clinical data from EHR and claims, which we specifically refer to here in this paper as CFM. CFM are mainly built to model a patient’s longitudinal medical history, condensing structured data, such as diagnoses, medications, procedures, and other structured data, into compact patient embeddings for predictive tasks.

**Med-BERT:** Med-BERT^3^ is an early encoder-based CFM developed to process structured electronic EHR data for predictive modeling tasks in healthcare. It applies the Transformer-based architecture of Bidirectional Encoder Representations from Transformers (BERT) to sequential EHR data, where medical codes like diagnoses are embedded and processed in temporal order. Med-BERT is pre-trained on large-scale clinical datasets using masked language modeling, where portions of patient records are masked, and the model learns to predict the missing information. By fine-tuning its pre-trained representations on task-specific datasets, Med-BERT has demonstrated better performance compared to traditional machine learning models and deep learning models without Med-BERT in two downstream tasks DHF and PaCa prediction.

**CLMBR:** CLMBR is a decoder transformer-based, autoregressive foundation model (CLMBR-T-base) originally described by Steinberg et al. (2021)^20^ and improved by Wornow et al. (2023)^19^. It is pretrained on 2.57 million de-identified EHRs from Stanford Medicine, using an objective that predicts the next medical code in a patient’s timeline given previous codes. Unlike BERT-based models which are bidirectional in nature, CLMBR-T-base employs forward-only, causally masked local attention to ensure that information only flows in one direction for predictive modeling. The model handles structured EHR data at minute-level granularity (rather than aggregating codes at the day level), and has 141 million parameters with a hidden dimension of 768. It does not process clinical text, but instead focuses only on structured information, allowing for robust patient representations that can be applied to downstream clinical prediction tasks.

#### A2 Large Language Models (LLM)

Five open-source generalist LLMs were selected for fine-tuning: the Mistral-7B-Instruct-v0.2^21^ model and LLaMA^15,16^ series models, including LLaMA-2-13b-hf, LLaMA-3-8B-Instruct, LLaMA-3.1-8B-Instruct, and LLaMA-3.1-70B-Instruct. Mistral-7B is a 7-billion-parameter language model based on a transformer architecture that balances models delivering both high-level performance. One of the main techniques Mistral used is grouped-query attention (GQA)^22^, which is an attention mechanism designed to for faster inference by grouping related queries together. Mistral also includes a sliding-window attention (SWA)^23^ to improve its attention mechanism to focus on specific parts of the input sequence, thus handling longer sequences more effectively at a reduced computational cost. It has exhibited exceptional performance across various benchmarks, outperforming LLaMA-2-13b in different evaluations. The other series of LLMs we selected to test are LLaMA-2 and LLaMA-3. LLaMA-2 is an auto-regressive language model that has a standard Transformer architecture with extensions such as root mean square layer normalization (RMSNorm)^24^ for pre-normalization and swish-gated linear unit (SwiGLU)^25^ activation function. Additionally, it offered a context length of 4,096 tokens and was fine-tuned using reinforcement learning from human feedback (RLHF)^26^. In contrast, LLaMA-3 has several improvements over LLaMA-2 while keeping a similar basic structure of a decoder-only transformer setup, including a more efficient tokenizer with a vocabulary of 128K tokens for pre-training, leading to significantly enhanced model performance. To improve inference efficiency, LLaMA-3 uses GQA across both 8B and 70B sizes and trains models on sequences of the context length of 8,192 tokens. LLaMA-3.1 shares a similar model architecture with LLaMA-3, the main difference is a significantly longer context length of 128K tokens. The largest LLM we tested is LLaMA-3.1-70B-Instruct, and this decision was made to balance the speed and efficacy based on the computational resource.

Table S1 compares some of the key characteristics of the models we selected to evaluate in this study. Med-BERT pretraining, with only 17 million parameters, uses a 512-token context and an 82K-token vocabulary, and does not incorporate grouped-query attention (GQA). GQA is a mechanism that groups related queries together to improve context capture and reduce computational overhead. In contrast, the generalist LLMs are substantially larger (ranging from 7B to 70B parameters) with significantly longer context lengths (ranging from 4K to 128K tokens) and larger vocabularies (up to 128K tokens). It is worth noting that models such as Mistral, LLaMA-3, and LLaMA-3.1 employ GQA, which likely enhances their ability to manage long context dependencies.

**Table S1 Model comparison**

| **Model name** | **Parameters** | **Context length (tokens)** | **Vocabulary size (tokens)** | **GQA** |
| --- | --- | --- | --- | --- |
| Med-BERT pretraining | 17M | 512 | 82K | No |
| Mistral | 7B | 8K | 128K | Yes |
| LLaMA-2 | 13B | 4K | 32K | No |
| LLaMA-3 | 8B | 8K | 128K | Yes |
| LLaMA-3.1 | 8B/70B | 128K | 128K | Yes |
| Phi-4-mini-flash-reasoning | 4B | 64K | 200K | Yes |
| Medgemma-it | 4B/29B | 128K | 262K | Yes |
| Gte-Qwen2-7B-instruct | 8B | 131K | 151K | Yes |
| Gpt-oss-20b | 22B | 131K | 200K | Yes |
| DeepSeek-R1-Distill-Llama-70B | 71B | 128K | 128K | Yes |

Grouped-query attention (GQA) simplifies how LLMs understand large amounts of text by bundling similar pieces together. This makes the model faster and smarter, as it can focus on groups of words at a time instead of each word individually.

#### A3 Clinical Large Language Model (CLLM)

Me-LLaMA is a specialized CLLM developed for medical text analysis tasks and clinical diagnosis. It is built upon LLaMA-2 and trained through continual pre-training and instruction tuning using a large-scale dataset with 129B pre-training tokens, as well as an instruction tuning dataset with 214k samples. The dataset includes biomedical literature, clinical guidelines, and clinical notes for EHR to enrich the model’s understanding of medical contexts. Me-LLaMA outperforms existing open-source medical LLMs in zero-shot, few-shot and supervised learning settings. Additionally, it showed competitive results against GPT-4 for diagnosing complex clinical cases.

### Appendix B Model comparison

Table S2 presents the descriptive analysis of the generalist and clinical LLMs, highlighting key differences in their architectural configurations. Among the models, Mistral-7B-Instruct-v0.2 has the smallest total number of parameters, with 7.1 billion, while LLaMA-3.1-70B-Instruct is the largest model, containing 69.6 billion total parameters. Me-LLaMA, derived from LLaMA-2-13b, shares the same total number of parameters (12.9 billion) as LLaMA-2-13b-hf.

**Table S2 Descriptive analysis of the LLMs**

| **Model name** | **Number of total parameters** | **Trainable parameters (%)** |
| --- | --- | --- |
| Mistral-7B-Instruct-v0.2 | 7,124,307,968 | 13,639,680 (0.19%) |
| LLaMA-2-13b-hf | 12,878,259,200 | 26,224,640 (0.20%) |
| LLaMA-3-8B-Instruct | 7,518,572,544 | 13,639,680 (0.18%) |
| LLaMA-3.1-8B-Instruct | 7,518,572,544 | 13,639,680 (0.18%) |
| LLaMA-3.1-70B-Instruct | 69,568,602,112 | 65,552,384 (0.09%) |
| Me-LLaMA | 12,878,259,200 | 26,224,640 (0.20%) |

When considering trainable parameters, LLaMA-3.1-70B-Instruct has the largest number, with 65.6 million trainable parameters, although this represents only 0.09% of its total parameters, the smallest proportion among all the models analyzed. By contrast, Mistral-7B-Instruct-v0.2, LLaMA-3-8B-Instruct, and LLaMA-3.1-8B-Instruct each have 13.6 million trainable parameters, corresponding to 0.18%-0.19% of their total parameters. Similarly, Me-LLaMA and LLaMA-2-13b-hf both have 26.2 million trainable parameters, accounting for 0.20% of their respective total parameters, the highest percentage among the models. These differences in the size and proportion of trainable parameters suggest distinct trade-offs in computational efficiency and adaptability across the models. Larger models, such as LLaMA-3.1-70B-Instruct, offer the potential for capturing more nuanced patterns due to their size, but their relatively smaller percentage of trainable parameters may limit fine-tuning flexibility. On the other hand, smaller models like Mistral-7B-Instruct-v0.2 and LLaMA-3.1-8B-Instruct balance size and adaptability, making them potentially more efficient with limited computational resources.

### Appendix C Datasets and prediction tasks

This study focuses on two binary classification disease prediction tasks: DHF and PaCa, mirroring those addressed by Med-BERT. The objective of these tasks is to predict whether a patient will be diagnosed with DHF or PaCa based on their structured EHR history. The prediction relies on patient history, including past diagnoses, medications, procedures, and other information from the EHR.

#### C1 Med-BERT datasets

To ensure a fair comparison, we used the same datasets evaluated in the Med-BERT study. These include two datasets derived from a structured EHR database (DHF-EHR and PaCa-EHR), and a patient-level claims dataset, PaCa-Claims. DHF-EHR and PaCa-EHR focus on longitudinal patient data with detailed patient information including diagnosis, medication, and procedure. PaCa-Claims is a patient-level claims dataset that captures person-specific clinical utilization, expenditures, and enrollment across inpatient, outpatient, prescription drug, and carve-out services. Following Med-BERT’s methodology, all datasets were split into training, validation, and test sets in a 70:10:20 ratio for fine-tuning and evaluation.

#### C2 EHRSHOT PaCa dataset

To further expand the evaluation, we included the publicly available EHRSHOT dataset for the PaCa prediction task. Unlike the datasets utilized for Med-BERT earlier evaluation, EHRSHOT offers a smaller sample size from a single source but contains a higher number of unique codes per patient and a richer set of longitudinal data, with an average of 95 visits per patient. To explore the impact of data richness, we processed patient histories in three formats: (1) diagnosis-only includes only diagnosis information, which is the same as Med-BERT; (2) diagnosis, medication, and procedure, a richer representation similar to the Med-BERT v2 preprocessing; and (3) all-codes, which incorporates all available structured data, such as demographics and lab results.

### Appendix D Data preprocessing

#### D1 For Med-BERT

The data preprocessing of the EHR and claims datasets remains consistent with the method described in the Methods section of our previous Med-BERT publication. Patient records were first extracted from the structured data source, including diagnosis, medication, and procedure codes. Visit sequences were organized based on the time order, with each sequence truncated or padded to a fixed length to ensure uniform input sizes.

Using the EHRSHOT dataset, to ensure a fair test between Med-BERT and CLMBR, we first converted all patient information to the Med-BERT data format to identify any patients that needed to be removed due to missing data, such as missing visit IDs. After this step, we obtained a total of 3,810 pancreatic cancer-related patients. To maintain patient-level consistency, we set the prediction time for each patient to the maximum available value.

#### D2 For CLMBR

Once the datasets were finalized, we created three different test formats to evaluate CLMBR: (1) diagnosis-only, included data only from the condition_occurrence and visit_occurrence tables; (2) diagnosis, procedure, and medication, consisted of additional information from the procedure_occurrence and drug_exposure tables; (3) all-codes, using all available tables. Since the Med-BERT model lacks information other than diagnosis, medication, and procedure, the same set of 3,810 patients and their train-validation-test split (70:10:20) were then used for the all-codes format test. A small number of patients with no recorded events in condition_occurrence were removed to ensure data consistency.

For EHRSHOT data preparation to be used with CLMBR, we used scripts from the EHRSHOT GitHub repository. The first script generated the FEMR PatientDatabase, while the second script, which labeled patients, required modification to ensure only one row per patient, with prediction time set to the maximum available value. Additionally, since CLMBR requires predictions to be made one year before diagnosis, we removed any data from the preprocessed Med-BERT format dataset that was recorded within one year of the diagnosis date. After splitting the labeled patients into case and control groups, these data were used as inputs for Med-BERT. Finally, we ran the remaining preprocessing scripts from the GitHub repository without modifications, except for updating data paths.

#### D3 For Large Language Models Fine-tuning

Before fine-tuning, patient histories were preprocessed and converted from structured EHR into text formats suitable for input to LLMs. For the DHF-EHR, PaCa-EHR, and PaCa-Claims datasets, we only included diagnosis information. This preprocessing step follows the data selection used in CPLLM and LLaMA2-EHR, which ensures consistency with previous studies. For the EHRSHOT PaCa dataset, we expanded our preprocessing strategy by experimenting with three different variations of patient history representations. First, we prepared a "diagnosis only" version, which again included only diagnostic information for each patient. Second, we created a version including diagnoses, medications, and procedures. This approach matches the data structure used in Med-BERT v2. Finally, we tested an all-codes format, which includes all available EHR data in the patient history, such as demographics, lab results, and other relevant clinical features. For continuous variables such as lab results, we added the value and unit after each description. This preprocessing framework aimed to maximize the compatibility of structured EHR data with LLMs. Additional details on the preprocessing steps can be found in Figure S1 of the supplementary materials.

**Prompt design:** Prompt design was an important methodological decision in structuring patient histories for input to LLMs. We tested two prompt formats based on designs previously introduced in CPLLM and LLaMA2-EHR. Both formats convert structured EHR data into textual representations, but they differ in the level of detail provided. The CPLLM prompt presented diagnostic information in a straightforward textual sequence. In contrast, the LLaMA2-EHR prompt added an additional layer of information by including the frequency of each diagnosis throughout the patient’s history. This allowed us to evaluate the impact of incorporating event frequency on model performance. By testing these prompt formats, we aimed to balance the trade-off between information richness and input simplicity, ensuring optimal compatibility with the tested LLM architectures.

Figure S1 summarizes the data preprocessing framework. For the EHR and Claims datasets, structured data primarily consists of International Classification of Diseases (ICD) codes, whereas the EHRSHOT dataset uses Systematized Nomenclature of Medicine-Clinical Terms (SNOMED CT) codes. Following the Med-BERT preprocessing methods, CFMs convert the EHR data into lists that include all numerical encounter-level information for each patient. In contrast, LLM data preprocessing transforms the data into lists where each list comprises a label and a text string generated from one of two prompt formats: CPLLM or LLaMA2-EHR. The LLaMA2-EHR format includes frequency information for each medical code description, distinguishing it from the CPLLM format. This framework standardizes data representation for subsequent fine-tuning and evaluation steps.

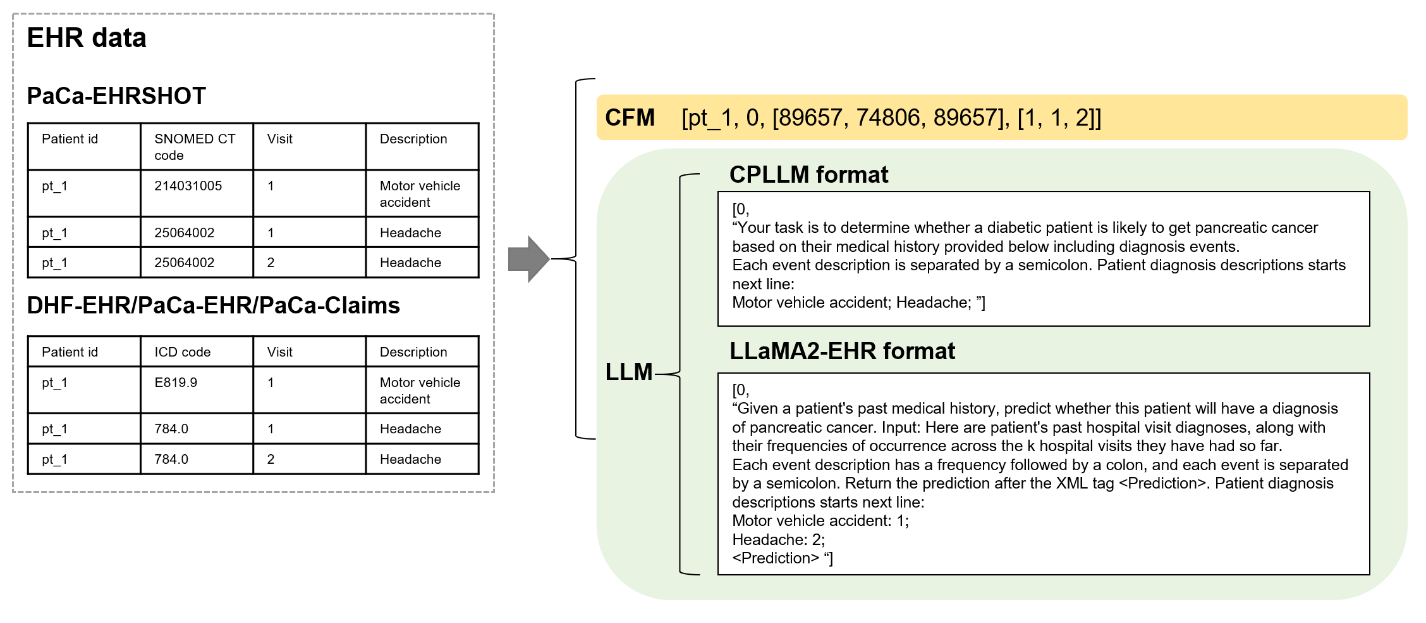

**Figure S1 Data preprocessing framework**

#### D4 Data preprocessing for LLM as Encoder

Utilizing predefined splits, each patient history is converted to an XML file and markdown file. Each file for a given patient was started with the patient’s demographic information: birth year, ethnicity, race and gender. For markdown files, a snapshot of the patient’s health was taken, which means that the most recently dated body metrics (e.g. body weight, body height), vitals (e.g. heart rate, blood pressure), risk factors (e.g. cigarette consumption), notable conditions (e.g. hyperlipidemia), and lab measurements (e.g. blood glucose) were outlined. After this summary, visits starting from most recent to oldest were outlined. Each visit contains diagnoses, medications, labs, and procedures that were assigned during the visit. XML formatted data reflects the structure of the markdown data without the patient summary in the beginning. Similar to the markdown files, patient visits were outlined from most recent to oldest. We then create 2 separate cohorts between both XML and markdown datasets: one cohort included an explicit instruction to predict each patient’s probability of developing PaCa, while the other cohort did not contain explicit instructions. These two cohorts, one with instructions and the other without, consisting of both XML and markdown formatted patient histories, was then fed into a frozen large language model. We use these frozen models to generate embeddings of the patient history. The last hidden layer is extracted, and we calculate the mean pooled embeddings from this layer. Finally, we train various classifiers on these embeddings and calculate the area under the receiver operating curve and area under the precision-recall curve.

### Appendix E Fine-tuning methods

#### E1 Clinical Foundation Models

**Frameworks and packages:** Both Med-BERT and CLMBR were implemented using PyTorch. Med-BERT fine-tuning used the HuggingFace Transformers library with the task set to sequence classification.

**Computational resources:** Both models were trained using high-performance GPU clusters. Med-BERT pretraining and fine-tuning were conducted on Nvidia V100 GPUs. CLMBR also required multi-GPU setups to efficiently process extensive longitudinal patient records, ensuring computational feasibility for large-scale representation learning.

**Hyperparameters:** For Med-BERT, the maximum sequence length was set to 512 tokens, and the model was trained using a batch size of 32 with the Adam optimizer with a learning rate of 1e-5. The pretraining was performed for a fixed number of epochs until convergence, with fine-tuning following a similar setup but with task-specific adjustments. CLMBR was trained with a maximum sequence length of 496 tokens and a learning rate of 1e-5. Batch sizes were adjusted based on GPU memory constraints, and fine-tuning involved optimizing patient embeddings with task-specific classifiers and applying early stopping criteria to prevent overfitting.

#### E2 Large Language Models / Clinical Large Language Model

**Frameworks and packages:** Fine-tuning was performed using the HuggingFace library with the task set to sequence classification. To enhance efficiency, we applied low-rank adaptation of large language models (LoRA)^33^ from the parameter-efficient fine-tuning (PEFT)^34^ techniques, which optimizes model performance while reducing computational overhead. Additionally, model quantization was implemented using the bitsandbytes library to decrease memory usage and accelerate model execution while maintaining accuracy.

**Computational resources:** All LLM fine-tuning experiments were performed using Nvidia H100 GPUs. Most fine-tuning tasks required two GPUs per run. However, fine-tuning LLaMA-3.1-70B-Instruct on the EHRSHOT dataset using the all-codes format required four GPUs per run due to the increased computational demand.

**Hyperparameters:** The maximum sequence length was set to 4,096 or 8,192 tokens, depending on the dataset and task requirements. For standard fine-tuning runs requiring 2 GPUs, the training batch size per GPU was set to 32, and the evaluation batch size per GPU was 16. For experiments requiring 4 GPUs, the batch sizes were adjusted to maintain consistency, with a training batch size per GPU of 16 and an evaluation batch size per GPU of 8. The learning rate was set as 2e-5 for all tasks, and the optimizer was Adam W (PyTorch). Hyperparameters related to LoRA were set the same as CPLLM.

### Appendix F Evaluation and experiments

Following the Med-BERT paper and other previous works on clinical prediction tasks, we used the area under the receiver operating characteristic curve (AUROC) as the evaluation metric for all experiments. For the DHF-EHR, PaCa-EHR, and PaCa-Claims tasks, which are the same as those in Med-BERT, we used diagnosis information as input, evaluated three traditional machine learning models: logistic regression (LR), random forest (RF), and light gradient boosting machine (LGBM). Med-BERT was the only clinical CFM tested for these tasks. Additionally, all five generalist LLMs and the CLLM were fine-tuned and evaluated on these three tasks.

For the PaCa task on EHRSHOT, we examined three different data formats: (1) diagnosis-only, which includes only diagnosis information; (2) diagnosis, medication, and procedure, following the format used in Med-BERT v2; and (3) all codes, which includes all available structured data in the dataset. The same three traditional machine learning models were evaluated, along with two CFMs, CLMBR and Med-BERT v2. Unlike the previous tasks, only the two best-performing LLMs from the DHF-EHR, PaCa-EHR, and PaCa-Claims experiments were selected for fine-tuning on the PaCa-EHRSHOT dataset. The same CLLM was also fine-tuned and evaluated on this task.

### Appendix G Descriptive analysis

Compared to the EHR (60,000 and 31,243 patients) and claims (33,850 patients) datasets, the EHRSHOT dataset contains fewer patients (3,810), but it provides a significantly higher average number of visits per patient (95) than the other datasets. This suggests that although the EHRSHOT dataset has a smaller sample size, it captures more comprehensive longitudinal information per patient. Additionally, the EHRSHOT dataset includes a greater number of unique codes and a higher average number of codes per patient compared to the EHR and claims datasets.

In terms of tokenization for LLM-based modeling, the DHF-EHR task contains the highest number of unique LLM tokens across all models, whereas the PaCa-EHRSHOT task with diag_only scope has the smallest number of unique LLM tokens. However, when considering average LLM tokens per patient, the PaCa-EHRSHOT task with all_codes scope achieves the largest value across all models, reflecting the detailed and extensive patient information captured in this dataset. In contrast, the PaCa-EHR task has the smallest average number of LLM tokens per patient due to its relatively smaller scope and sample size. These findings illustrate the variation in dataset composition and task complexity, providing critical context for interpreting the model performance results.

### Appendix H Fine-tuning results

Table S3 presents the average AUC and standard deviations over 3 repeats for three fine-tuned LLMs using the CPLLM prompt format on the prediction tasks using the EHR and Claims datasets (DHF-EHR, PaCa-EHR, and PaCa-Claims). On the DHF-EHR task, the models show comparable performance with AUROCs of 83.33 (Mistral-7B-Instruct-v0.2), 83.57 (LLaMA-2-13b-hf), and 83.42 (LLaMA-3-8B-Instruct). For the PaCa-EHR task, LLaMA-2-13b-hf leads with an average AUROC of 82.65, followed by LLaMA-3-8B-Instruct at 82.07 and Mistral-7B-Instruct-v0.2 at 81.88. In the PaCa-Claims task, the performance slightly declines, with LLaMA-3-8B-Instruct achieving 79.53, LLaMA-2-13b-hf 79.41, and Mistral-7B-Instruct-v0.2 79.25.

**Table S3 Model performance for fine-tuned LLMs based on the CPLLM prompt format**

| **Model name** | **Average AUROC (STD)** | | |
| --- | --- | --- | --- |
|  | **DHF-EHR** | **PaCa-EHR** | **PaCa-Claims** |
| Mistral-7B-Instruct-v0.2 | 83.33 (0.44) | 81.88 (0.17) | 79.25 (0.12) |
| LLaMA-2-13b-hf | 83.57 (0.08) | 82.65 (0.12) | 79.41 (0.29) |
| LLaMA-3-8B-Instruct | 83.42 (0.16) | 82.07 (0.12) | 79.53 (0.22) |

AUROC: the area under the receiver operating characteristics curve; STD: standard deviation for 3 repeats.

The ML and CFM results for the DHF-EHR, PaCa-EHR, and PaCa-Claims tasks are copied from the results table of the Med-BERT paper. For the DHF-EHR task, the CFM Med-BERT + Bi-GRU achieved the highest AUROC of 85.39 (0.05), outperforming other models, while Me-LLAMA and LLaMA-3.1-70B-Instruct were the leading LLMs with comparable AUROCs of 84.46 (0.06) and 84.73 (0.20), respectively. For PaCa-EHR, the LLaMA-3.1-70B-Instruct model and Me-LLAMA demonstrated superior performance with AUROCs of 82.96 (0.16) and 82.96 (0.30), respectively, narrowly surpassing Med-BERT + Bi-GRU’s 82.23 (0.29). It is worth noting that Med-BERT + Bi-GRU excelled in the PaCa-Claims task, with an AUROC of 80.57 (0.21), outperforming both generalist LLMs and clinical LLMs, such as Me-LLAMA, which achieved 79.87 (0.15). Interestingly, traditional ML models such as logistic regression (LR) and random forest (RF) generally exhibit lower AUROCs than CFMs and LLMs, highlighting the superior predictive capabilities of advanced models. The p-value comparisons between CFMs and the best-performing LLMs are significant across all tasks, emphasizing that CFMs can match or outperform fine-tuned LLMs in these scenarios.

We tested model performance for the PaCa-EHRSHOT task under three scopes of patient information: diag_only (only include the diagnosis information), diag+med+proc (include diagnosis, medication, and procedure information), and all_codes (include all available information). Since the original Med-BERT model was pre-trained only using the diagnosis information, we selected to test Med-BERT v2, which was pre-trained on diagnosis, medication, and procedure information. Additionally, we only tested the better prompt format and the best generalist LLM based on the previous experiments, which are the LLaMA2-EHR prompt format and the LLaMA-3.1 series models. The results indicate that the LLaMA-3.1-70B-Instruct model achieved the highest AUROC of 86.1 (0.94) in the diag_only setting, while the best result from CFMs is the AUROC of 85.25 from Med-BERT v2 + Bi-GRU. Under the diag+med+proc format, LLaMA-3.1-70B-Instruct again excelled with an AUROC of 86.65 (1.19), followed closely by CLMBR at 86.33 (0). For the all_codes format, LGBM achieved the best performance with an AUROC of 90.26 (0.02), slightly outperforming LR at 88.37 (0.03). CFMs exhibited competitive performance in specific settings, such as Med-BERT v2 + Bi-GRU achieving an AUROC of 85.25 (1.30) in diag_only and CLMBR excelling in diag+med+proc with an AUROC of 86.33 (0). However, CLMBR didn’t achieve competitive results in broader scopes like all_codes, where data complexity increases. Additionally, the performance of traditional ML models showed AUROC values significantly lower than both CFMs and LLMs for the diag_only and diag+med+proc formats, but provided higher AUROC for the all_codes format when compared with both CFMs and LLMs. We calculated p-values using an unpaired t-test to compare the difference between the best CFM and the best LLM. The all_codes format is the only data format that makes a significant difference between the model performance of the CFM and the best LLM (p=0.0043).

Additionally, we reported the secondary results using AUPRC based on the EHRSHOT dataset tasks. The results show that even for tasks where CFMs and traditional ML models provided lower AUROCs than LLMs, they can still achieve higher AUPRCs. For the diag_only format, Med-BERT v2 + Bi-GRU achieved the highest AUPRC of 55.85 (3.99), followed by Med-BERT v2 with an AUPRC of 46.36 (5.16). In comparison, LLMs showed lower AUPRC values with relatively higher standard deviations, with LLaMA-3.1-8B-Instruct achieving 40.57 (5.47) and LLaMA-3.1-70B-Instruct achieving 41.14 (3.9). For the diag+med+proc format, CLMBR is the CFM that achieved the highest AUPRC of 54.9 (0), while Med-BERT v2 + Bi-GRU provided the second-highest AUPRC of 54.04 (7.81). Similar to the results of the diag_only format, both LLMs provided lower AUPRC than all CFMs, with LLaMA-3.1-8B-Instruct at 34.81 (12.46) and LLaMA-3.1-70B-Instruct at 49.07 (1.83). In the all_codes format, consistent with the AUROC results, the traditional ML model LGBM achieved the highest AUPRC of 51.95 (0.08). Among CFMs and LLMs, CLMBR provided a higher AUPRC of 48.25 (0), while LLaMA-3.1-8B-Instruct and LLaMA-3.1-70B-Instruct achieved lower AUPRCs of 37.03 (9.09) and 46.64 (7), respectively.

### Appendix I Computational resources comparison

Moreover, Tables S4 and S5 compare the fine-tuning process speed by hours for all generalist and clinical LLMs. Among the LLMs fine-tuned using the Med-BERT evaluation datasets, Mistral-7B-Instruct-v0.2 demonstrated the fastest fine-tuning speed, particularly on the DHF-EHR dataset (42 hours), while the largest model, LLaMA-3.1-70B-Instruct, required significantly more time, up to 190 hours. The fine-tuning process takes less time on the small dataset EHRSHOT. LLaMA-3.1-8B-Instruct achieved the fastest results for diagnostic codes only (3 hours), while LLaMA-3.1-70B-Instruct demands up to 110 hours for the all-codes setting. In contrast, CFMs require much less fine-tuning time when compared with LLMs, given that both Med-BERT and CLMBR require less than 1 hour, even for the task that the largest LLM requires 190 hours.

**Table S4 Speed comparison for fine-tuning LLMs using the EHR and Claims datasets**

| **Model name** | **Fine-tuning speed (hours)** | | |
| --- | --- | --- | --- |
|  | **DHF-EHR** | **PaCa-EHR** | **PaCa-Claims** |
| Mistral-7B-Instruct-v0.2 | 42 | 13 | 18 |
| LLaMA-2-13b-hf | 80 | 24 | 33 |
| LLaMA-3-8B-Instruct | 33 | 11 | 14 |
| LLaMA-3.1-8B-Instruct | 33 | 11 | 14 |
| LLaMA-3.1-70B-Instruct | 190 | 65 | 85 |
| Me-LLaMA | 80 | 25 | 35 |

**Table S5 Speed comparison for fine-tuning LLMs using the EHRSHOT dataset**

| **Model name** | **Fine-tuning speed (hours) for PaCa-EHRSHOT** | | |
| --- | --- | --- | --- |
|  | **D** | **DMP** | **ALL** |
| LLaMA-3.1-8B-Instruct | 3 | 4 | 19 |
| LLaMA-3.1-70B-Instruct | 16 | 18 | 110 |
| Me-LLaMA | 5 | 7 | 30 |

### Appendix J LLM as encoder additional results

In addition to finetuning Llama-3.1-70B-Instruct, we explored additional methodologies to compare predictive performance while attempting to be efficient with computing resources. Using all patient codes and our existing PaCa cohort, we first format patient histories in either XML, markdown, or summary text format, and proceeded to generate embeddings for said histories. By feeding these embeddings into binary classifiers, we measure performance with AUROC and AUPRC and Table S6 compares these results with our finetuning results for Llama-3.1-70B-Instruct.

**Table S6 Llama-3.1-70B-Instruct comparison against different methods and data formats**

| **Llama 3.1 70B** | **AUROC** | | **AUPRC** | |
| --- | --- | --- | --- | --- |
| **Text Format** | **Fine-tuning** | **No Finetuning** | **Fine-tuning** | **No Finetuning** |
|  | 84.7 | 87.0 | 46.6 | 50.6 |
| **Additional File Format - with No Fine- tuning** | **XML** | **Markdown** | **XML** | **Markdown** |
|  | 83.4 | **88.5** | 37.0 | **59.9** |

AUROC: the area under the receiver operating characteristics curve; AUPRC: the area under the precision-recall curve. Bold numbers are the best AUROC or AUPRC.

Tables S7 and S8 showing the results for LLM as encoders without and with instructions, separately.

**Table S7 LLM as encoders results for cohort without instructions**

|  |  | **Phi 4B** | **MedGemma 4B** | **Qwen2 7B** | **GPT OSS 20B** | **MedGemma 27B** | **Llama 3.1 70B** | **DeepSeek R1 Distill 70B** |
| --- | --- | --- | --- | --- | --- | --- | --- | --- |
|  |  | **AUROC (STD)** | | | | | | |
| **XML** | LR | 0.800 (0.000) | 0.826 (0.000) | 0.828 (0.000) | 0.799 (0.000) | 0.829 (0.000) | 0.833 (0.000) | 0.842 (0.000) |
|  | Feed Forward NN | 0.801 (0.003) | 0.821 (0.003) | 0.837 (0.006) | 0.788 (0.023) | 0.837 (0.005) | 0.815 (0.005) | 0.821 (0.005) |
|  | XGBoost | 0.625 (0.000) | 0.726 (0.000) | 0.724 (0.000) | 0.669 (0.000) | 0.682 (0.000) | 0.734 (0.000) | 0.679 (0.000) |
|  | RF | 0.692 (0.016) | 0.707 (0.006) | 0.719 (0.010) | 0.706 (0.012) | 0.716 (0.013) | 0.692 (0.011) | 0.689 (0.008) |
|  | LGBM | 0.645 (0.000) | 0.722 (0.000) | 0.739 (0.000) | 0.730 (0.000) | 0.692 (0.000) | 0.721 (0.000) | 0.746 (0.000) |
| **Markdown** | LR | 0.853 (0.000) | 0.856 (0.000) | 0.849 (0.000) | 0.881 (0.000) | 0.868 (0.000) | **0.885 (0.000)** | **0.885 (0.000)** |
|  | Feed Forward NN | 0.861 (0.004) | 0.863 (0.003) | 0.868 (0.008) | 0.862 (0.004) | 0.865 (0.006) | 0.867 (0.005) | 0.867 (0.007) |
|  | XGBoost | 0.767 (0.000) | 0.732 (0.000) | 0.771 (0.000) | 0.751 (0.000) | 0.809 (0.000) | 0.749 (0.000) | 0.798 (0.000) |
|  | RF | 0.766 (0.009) | 0.763 (0.009) | 0.790 (0.011) | 0.779 (0.009) | 0.759 (0.013) | 0.751 (0.018) | 0.759 (0.008) |
|  | LGBM | 0.763 (0.000) | 0.752 (0.000) | 0.764 (0.000) | 0.775 (0.000) | 0.790 (0.000) | 0.716 (0.000) | 0.822 (0.000) |
|  |  | **AUPRC (STD)** | | | | | | |
| **XML** | LR | 0.330 (0.000) | 0.376 (0.000) | 0.348 (0.000) | 0.309 (0.000) | 0.336 (0.000) | 0.365 (0.000) | 0.351 (0.000) |
|  | Feed Forward NN | 0.332 (0.014) | 0.317 (0.017) | 0.363 (0.022) | 0.256 (0.040) | 0.385 (0.009) | 0.354 (0.015) | 0.361 (0.024) |
|  | XGBoost | 0.172 (0.000) | 0.184 (0.000) | 0.208 (0.000) | 0.209 (0.000) | 0.134 (0.000) | 0.262 (0.000) | 0.174 (0.000) |
|  | RF | 0.218 (0.009) | 0.215 (0.026) | 0.191 (0.016) | 0.228 (0.010) | 0.220 (0.024) | 0.195 (0.004) | 0.192 (0.015) |
|  | LGBM | 0.127 (0.000) | 0.199 (0.000) | 0.224 (0.000) | 0.237 (0.000) | 0.169 (0.000) | 0.233 (0.000) | 0.241 (0.000) |
| **Markdown** | LR | 0.469 (0.000) | 0.549 (0.000) | 0.513 (0.000) | **0.650 (0.000)** | 0.562 (0.000) | 0.581 (0.000) | 0.594 (0.000) |
|  | Feed Forward NN | 0.471 (0.024) | 0.501 (0.031) | 0.594 (0.033) | 0.494 (0.050) | 0.542 (0.015) | 0.480 (0.036) | 0.518 (0.025) |
|  | XGBoost | 0.254 (0.000) | 0.253 (0.000) | 0.355 (0.000) | 0.260 (0.000) | 0.450 (0.000) | 0.211 (0.000) | 0.321 (0.000) |
|  | RF | 0.227 (0.012) | 0.280 (0.021) | 0.309 (0.017) | 0.257 (0.014) | 0.257 (0.021) | 0.215 (0.025) | 0.221 (0.012) |
|  | LGBM | 0.335 (0.000) | 0.296 (0.000) | 0.272 (0.000) | 0.259 (0.000) | 0.257 (0.000) | 0.215 (0.000) | 0.320 (0.000) |

AUROC: the area under the receiver operating characteristics curve; AUPRC: the area under the precision-recall curve; STD: standard deviation for 3 repeats.

Bold numbers are the best average AUROC or AUPRC.

**Table S8 LLM as encoders results for cohort with instructions**

|  |  | **Phi 4B** | **MedGemma 4B** | **Qwen2 7B** | **GPT OSS 20B** | **MedGemma 27B** | **Llama 3.1 70B** | **DeepSeek R1 Distill 70B** |
| --- | --- | --- | --- | --- | --- | --- | --- | --- |
|  |  | **AUROC (STD)** | | | | | | |
| **XML** | LR | 0.798 (0.000) | 0.823 (0.000) | 0.827 (0.000) | 0.839 (0.000) | 0.825 (0.000) | 0.834 (0.000) | 0.852 (0.000) |
|  | Feed Forward NN | 0.803 (0.004) | 0.824 (0.004) | 0.832 (0.006) | 0.828 (0.016) | 0.841 (0.002) | 0.820 (0.008) | 0.831 (0.006) |
|  | XGBoost | 0.694 (0.000) | 0.675 (0.000) | 0.707 (0.000) | 0.766 (0.000) | 0.715 (0.000) | 0.701 (0.000) | 0.697 (0.000) |
|  | RF | 0.683 (0.009) | 0.698 (0.015) | 0.698 (0.012) | 0.745 (0.009) | 0.724 (0.007) | 0.702 (0.014) | 0.701 (0.015) |
|  | LGBM | 0.690 (0.000) | 0.648 (0.000) | 0.698 (0.000) | 0.736 (0.000) | 0.711 (0.000) | 0.677 (0.000) | 0.788 (0.000) |
| **Markdown** | LR | 0.846 (0.000) | 0.858 (0.000) | 0.849 (0.000) | 0.884 (0.000) | 0.869 (0.000) | 0.885 (0.000) | **0.899 (0.000)** |
|  | Feed Forward NN | 0.851 (0.005) | 0.860 (0.002) | 0.860 (0.013) | 0.866 (0.004) | 0.872 (0.006) | 0.866 (0.007) | 0.871 (0.005) |
|  | XGBoost | 0.793 (0.000) | 0.753 (0.000) | 0.746 (0.000) | 0.829 (0.000) | 0.818 (0.000) | 0.802 (0.000) | 0.703 (0.000) |
|  | RF | 0.758 (0.010) | 0.765 (0.012) | 0.751 (0.005) | 0.825 (0.011) | 0.769 (0.010) | 0.749 (0.011) | 0.763 (0.008) |
|  | LGBM | 0.776 (0.000) | 0.785 (0.000) | 0.721 (0.000) | 0.806 (0.000) | 0.799 (0.000) | 0.766 (0.000) | 0.712 (0.000) |
|  |  | **AUPRC (STD)** | | | | | | |
| **XML** | LR | 0.331 (0.000) | 0.329 (0.000) | 0.341 (0.000) | 0.430 (0.000) | 0.317 (0.000) | 0.371 (0.000) | 0.374 (0.000) |
|  | Feed Forward NN | 0.350 (0.021) | 0.325 (0.019) | 0.363 (0.017) | 0.372 (0.041) | 0.403 (0.019) | 0.387 (0.015) | 0.379 (0.026) |
|  | XGBoost | 0.240 (0.000) | 0.213 (0.000) | 0.156 (0.000) | 0.343 (0.000) | 0.262 (0.000) | 0.177 (0.000) | 0.203 (0.000) |
|  | RF | 0.218 (0.017) | 0.234 (0.026) | 0.184 (0.011) | 0.305 (0.010) | 0.242 (0.016) | 0.181 (0.012) | 0.201 (0.014) |
|  | LGBM | 0.224 (0.000) | 0.174 (0.000) | 0.139 (0.000) | 0.206 (0.000) | 0.201 (0.000) | 0.202 (0.000) | 0.320 (0.000) |
| **Markdown** | LR | 0.463 (0.000) | 0.549 (0.000) | 0.534 (0.000) | **0.661 (0.000)** | 0.594 (0.000) | 0.599 (0.000) | 0.599 (0.000) |
|  | Feed Forward NN | 0.460 (0.021) | 0.489 (0.020) | 0.536 (0.037) | 0.540 (0.035) | 0.579 (0.018) | 0.517 (0.036) | 0.539 (0.034) |
|  | XGBoost | 0.363 (0.000) | 0.358 (0.000) | 0.266 (0.000) | 0.419 (0.000) | 0.346 (0.000) | 0.295 (0.000) | 0.168 (0.000) |
|  | RF | 0.245 (0.010) | 0.276 (0.016) | 0.257 (0.020) | 0.399 (0.006) | 0.256 (0.015) | 0.198 (0.011) | 0.225 (0.007) |
|  | LGBM | 0.275 (0.000) | 0.356 (0.000) | 0.338 (0.000) | 0.402 (0.000) | 0.309 (0.000) | 0.211 (0.000) | 0.188 (0.000) |

AUROC: the area under the receiver operating characteristics curve; AUPRC: the area under the precision-recall curve; STD: standard deviation for 3 repeats.

Bold numbers are the best average AUROC or AUPRC.
